# Beyond prevention: A rightward shift in the distribution of weight-for-height following supplementation with SQ-LNS to children aged 6–23 months in post-conflict settings of Tigray, Ethiopia - A Non-Randomized Cluster Trial

**DOI:** 10.64898/2026.03.30.26349799

**Authors:** Afework Mulugeta Bezabih, Ramadhani Noor, Meseret Demissie, Gebretsadkan Gebremedhin Gebretsadik, Hadush Gebregziabher, Kidanemariam Alem, Mulugeta Woldu, Leela Zayzay, Yosef Teklu, Yemane Hailu, Nigsti Tsegay, Tesfay Gebreegziabher, Rieye Esayas, Mengish Bahresellasie, Ashenafi Asmelash, Hailay Kidane, Dawit Seyoum, Stanley Chitekwe

## Abstract

**Introduction:** Acute malnutrition in children aged 6–23 months remains critical in Tigray, Ethiopia, where global acute malnutrition (GAM) rates have reached emergency levels. Small-quantity lipid-based nutrient supplements (SQ-LNS) show promise for prevention, but evidence from post-conflict settings is limited.

**Objective:** This study evaluated SQ-LNS effectiveness in preventing acute malnutrition and rightward shifting in the distribution of weight-for-height among young children in post-conflict settings of Tigray, Ethiopia.

**Methods:** A non-randomized cluster trial enrolled 8,145 children aged 6–23 months across four districts. The intervention group (n=6,752) received daily 20g SQ-LNS sachets for six months plus behavior change communication; the control group (n=1,393) received standard nutrition programming. Primary outcomes were acute malnutrition prevalence (WHZ < -2 or MUAC < 12.5cm) and distribution of weight-for-height z-scores. Data were collected biweekly and analyzed using longitudinal comparisons and difference-in-differences (DiD) estimation.

**Results:** Acute malnutrition declined from 22.2% to 4.6% in the intervention group (17.6 percentage point reduction) versus 20.1% to 14.0% in controls (6.1-point reduction). Mean WHZ scores increased from -0.35 to +0.37 in the intervention group (gain of +0.72 z-scores), while controls improved from -0.79 to -0.63 (gain of +0.16). The net intervention effect (DiD) showed a 4.1 percentage point reduction in WHZ-defined GAM and a 11.5-point reduction in MUAC-defined GAM. Mean WHZ and MUAC increased significantly more in the intervention group (DiD: +0.56 z-scores and +5.03mm, respectively). Critically, the entire WHZ distribution shifted rightward, indicating population-level nutritional improvement, not merely reduced caseloads.

**Conclusions:** Six months of daily SQ-LNS effectively prevented acute malnutrition and shifted the entire weight-for-height distribution rightward among young children in post-conflict settings of Tigray, Ethiopia. Benefits extended beyond treatment, lifting the whole child population’s nutritional status. Findings support SQ-LNS inclusion in post-conflict nutrition packages and highlight the importance of assessing distributional outcomes, not just prevalence, when evaluating nutritional interventions.

**Trial registration number:** This trial was registered at ClinicalTrials.gov, NCT06103084.

## Introduction

Malnutrition continues to affect millions of people in areas where extreme poverty and food insecurity remain deeply entrenched [1]. Globally, an estimated 149 million children under the age of 5 years were suffering from stunting in 2022 [2], and 47 million were wasted in 2019 [3]. Moreover, it is projected that 582 million people will be chronically undernourished at the end of the decade, by 2030, more than half of them in Africa [1]. Similarly, the burden of both acute and chronic malnutrition among children under five in Ethiopia remains high [4]. Nationally, 41% and 11% of Ethiopian children under five years of age are stunted and wasted, respectively [4]. Only 8% of the children satisfied the minimum dietary diversity (≥ 5 food groups) and 7% fulfilled the minimum acceptable diet (MAD) [4]. Moreover, child food poverty was a severe public health problem in Ethiopia. About half (47%) of children under five suffered from severe food poverty (consumed < 2 food groups a day), and about 45% of the children aged 6-23 months had moderate food poverty (consumed 3-4 food groups a day) [4]. Moreover, Ethiopian children under five are characterized by the highest burden of zero vegetable or fruit consumption (74.0%), suggesting that the largest proportion of the children were unable to afford a healthy diet [4].

The malnutrition situation in Ethiopia reflects where progress has been made, but not robust enough to alter the fundamental trajectory for millions of children. A continuation of the past trends simply means that millions of children will still be affected by malnutrition in its different forms, and the country will still be falling short of filling the nutrient gaps and reaching the global nutrition targets. Despite these efforts, fundamental challenges persist around child malnutrition. Systemic weaknesses in the health systems, agriculture, and social protection continue to limit the reach and effectiveness of nutrition interventions. Moreover, the monotony of Ethiopian children’s diets, with only 8% meeting minimum dietary diversity, represents a critical failure point that requires attention to food systems, affordability, and behavior change [4].

One promising approach to bridging this nutrient gap, particularly during the critical complementary feeding period when dietary diversity is most deficient, is the use of specially formulated nutrient supplements. Amongst these supplements are the small-quantity lipid-based nutrient supplements (SQ-LNS), which are developed to address nutritional deficiencies, including gaps in the key nutrients required for growth [5, 6]. The 20-gram SQ-LNS provides energy, protein, essential fatty acids, and a range of vitamins and minerals [7]. The dose of the SQ-LNS is kept as small as possible in order to avoid displacement of breastmilk and locally available nutrient-rich foods [8]. The evidence base for the preventive effect of SQ-LNS on acute malnutrition during the complementary feeding period (6–23 months) has been reported in various studies [8, 9, 10]. Children who received SQ-LNS had a lower prevalence of stunting, wasting, and underweight than children who did not [5, 7, 9, 11].

Given the persistent high rates of malnutrition and poor dietary diversity in Ethiopia, coupled with the growing global evidence, there is a clear rationale for exploring the scalability of SQ-LNS as a preventive intervention within the national nutrition program. However, the preventive effects of daily SQ-LNS during the first two years of life in post conflict settings are less documented in the Ethiopian setting, though the prevalence of acute malnutrition among these children has remained persistently high without a declining trend despite the implementation of several nutrition interventions [4, 12–14].

Although recent commitments to increase funding for the early detection and treatment of child wasting are encouraging, the prevention of wasting using proven interventions should also be prioritized in Ethiopia. The present study is unique in the sense that it is conducted in post conflict settings where the disruption and displacement of populations in emergencies pose an added threat to the existing situation of acute malnutrition among 6 – 23 months old children. In the present study, we reported the results of a follow-up evaluation of acute malnutrition among children who were supplemented with SQ-LNS during the complementary feeding period in post conflict settings. Our findings suggested that SQ-LNS supplementation brought a rightward shift of the distribution of the nutritional status and a protective effect on acute malnutrition among 6 – 23 months old children from post conflict settings, where the prevalence of child malnutrition was high. We hypothesized that children in the SQ-LNS group would have a lower burden of acute malnutrition compared to the children in the existing nutrition programs (the control group).

## Methodology

### Study setting

This trial was conducted in three purposively selected intervention districts, namely Enda Mekhoni, Maichew, and Neqsege, from the southern zone of Tigray, and one control district known as Ahse’a, from the central zone of Tigray, Northern Ethiopia. The intervention and control districts were the project areas of Tigray Development Association (TDA) and Mums for Mums (M4Ms), respectively. Maichew is a town and district located about 665 km north of Addis Ababa, the Capital of Ethiopia. The district is classified under the Weinadega (semi-temperate zone) and has a population of around 23,419 people. Enda Mekhoni district has a population of approximately 84,739 people, with the majority being Tigrayan and practicing Ethiopian Orthodox Christianity. Neqsege is a mountainous district that has a total population of 17,578. The control district, Ahse’a, is located in the central zone of Tigray. Agriculture is the mainstay of the three rural districts, namely Enda Mekhoni, Neqsege, and Ahse’a districts. The study was conducted from July 2023 to April 2024.

### Study design

The study was originally designed as a three-arm non-randomized cluster trial comparing: (i) Bahgina intervention, (ii) SQLNS intervention, and (iii) a control group. The target population for all three arms was children aged 6–23 months. However, during implementation, resource constraints necessitated a modification to the Bahgina intervention arm, which was delivered only to children aged 6–11 months. In contrast, the SQLNS and control arms were implemented as originally planned for children aged 6–23 months. Consequently, the Bahgina arm was no longer comparable to the other two arms with respect to age eligibility. Therefore, this manuscript reports findings from the SQLNS and control arms only, both of which retained the original age criteria of 6–23 months. The Bahgina arm is not included in the present analysis. In the non-randomized cluster trial, districts were assigned to either the intervention or the control group through non-random means. A non-randomized rollout of SQ-LNS was implemented alongside existing community-based nutrition interventions in Maichew, Enda Mekhoni, and Neqsege districts against Ahse’a, the control district with existing community-based nutrition interventions. The study protocol was registered at ClinicalTrials.gov with the trial number NCT06103084 and trial URL: https://clinicaltrials.gov/.

### Study population and eligibility criteria

The study population included children aged 6 – 23 months residing in Enda Mekhoni, Maichew, Neqsege, and Ahse’a districts. All children aged 6 – 23 months from the specified districts were included, and children with congenital defects, chronic diseases affecting growth and development, or known food allergies were excluded. This exclusion criterion was applied to minimize potential confounding factors and to ensure that the observed effects could be attributed more to the intervention. The transparent reporting of evaluations with a non-randomized design (TREND) flow diagram describing the participant recruitment and enrollment processes is shown in Figure 1.

**Figure 1:**
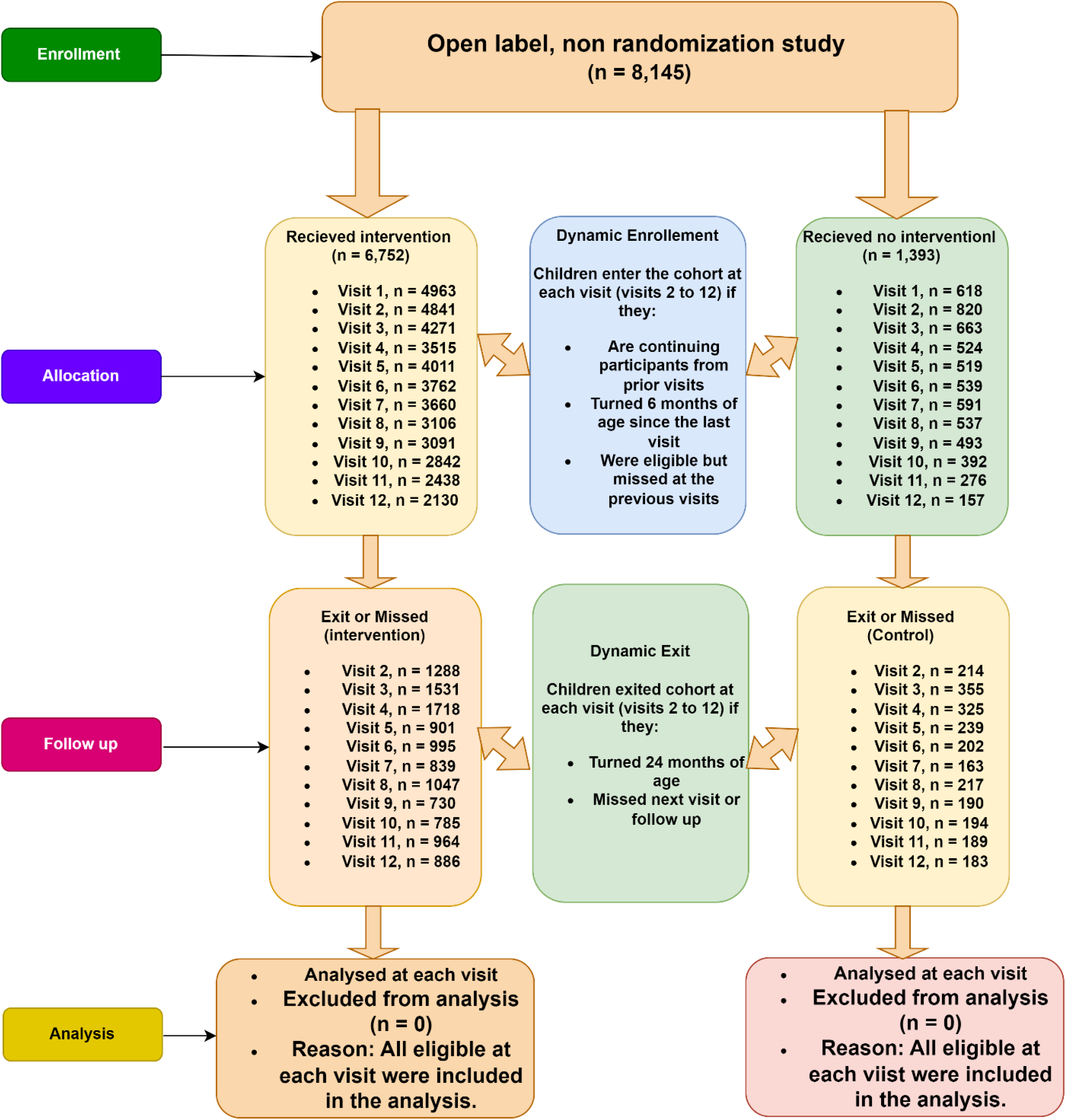
A flow diagram of participant progression through the study, as recommended for the transparent reporting of evaluations with non-randomized design (TREND) of the SQLNS follow-up study in the post-conflict settings of Tigray, Northern Ethiopia.

### Sample size determination and sampling technique

The sample size for this study was initially calculated based on a reported 6% and 22% respective prevalence of SAM and MAM in Tigray, Ethiopia [15], which resulted in 646 children from Maichew, 1,761 children from Enda Mekhoni, and 537 children from Neqsege, totaling 2,924 children in the intervention cluster. However, due to the very clear objective of the study being to determine the effectiveness of SQ – LNS in reducing the burden of acute malnutrition and due to the availability of enough SQ – LNS for the whole study period on a one-supplement per day per child calculation, all 6 – 23-month-old children in the three districts were included in the study. This amounted to 1991 children from Maichew, 3,396 children from Enda Mekhoni, and 1,365 children from Neqsege, resulting in a total intervention sample of 6752. Similarly, in the control district, all 6 – 23-month-old children (1393) were included in the study. The inclusion of the entire eligible population will ensure adequate statistical power and enhance external validity and generalizability of the findings.

### Intervention and intervention procedures

The six-month duration intervention was the provision of SQ – LNS to 6 – 23 months old children in the intervention cluster. The intervention was supplemented with the provision of key messages for children/caregivers regarding SQ – LNS and optimal infant and young child feeding (IYCF) practices by the health extension workers. The key messages provided included information on SQ – LNS’s beneficial effects for the well-being and development of children; the contraindication of SQ – LNS for infants under six months of age; the importance of not using it as a replacement for breastfeeding; the various methods of consumption, including direct ingestion from the sachet or mixing with other foods; its classification as a specialized supplementary food with optimal benefits when consumed consistently daily without sharing with family members; the necessity of seeking medical attention if children experience self-limiting diarrhea in the initial days of SQ – LNS consumption to exclude other potential causes of diarrhea, and optimal IYCF practices including timely introduction of solid, semi-solid or soft foods, minimum dietary diversity, minimum meal frequency, continued breastfeeding to two years and beyond and responsive feeding.

To run the intervention in a smoother way, a grassroots-level strategy was followed. The health system was the sole implementer of the intervention. Health extension workers (HEWs), Women Development Groups (WDGs), district health supervisors, health center directors, experts from the woreda health offices, and health centers were trained by the research team. The trainings of the HEWs and WDGs facilitated the identification and recruitment of 6 – 23 months old children into the SQ-LNS interventional study. It also resulted in community awareness and mobilization for the program and paved the way for an effective and smooth roll-out of the SQ-LNS intervention. Besides, monthly review meetings were held among all stakeholders in the intervention areas, which helped with timely tracking of challenges and their timely and appropriate addressing. The intervention was implemented at three levels: (i) “community level” to maximize identification and enrollment of eligible children, (ii) “facility level” for initiation of the interventions, and (iii) “household level” for maintenance and continuation of the interventions. For every 6 – 23-month-old child in the intervention districts, one sachet of SQ – LNS per day was provided for 6 months, which was equivalent to 180 sachets for a child with a complete follow-up. SQ – LNS sachets were provided biweekly at local health posts or health centers by trained HEWs or/and/or nurses.

### Outcomes

The outcome variable was the burden of acute malnutrition among 6 – 23-month-old children in the study communities from the post conflict settings in Tigray, Ethiopia. Acute malnutrition was defined as weight-for-length below -2 z scores and/or MUAC < 12.5 cm.

### Data collection tools and procedures

#### Follow-up data collection

Follow-up data were collected electronically via Open Data Kit (ODK) software every two weeks (fortnightly) using case-reporting forms (CRFs) The CRFs were used to gather data on SQ – LNS intake and anthropometric data. Nutrition-tailored messages were provided to the mothers or caregivers during each visit to increase adherence and compliance.

#### Anthropometric measurements

Anthropometric data were collected at baseline, and every two weeks throughout the six months of supplementation, following standard anthropometric procedures (11). Child’s recumbent length was measured to the nearest 0.1 cm by using a portable stadiometer with no footwear. Weight was measured by using the Electronic Digital Scale with the mother-child tare feature to the nearest 0.1 kg. Each child was weighed with minimal clothing and no footwear. MUAC was measured on the left upper arm at the midpoint between the acromion and olecranon process using non-stretchable tape recorded to the nearest 0.1cm. For all measurements, MUAC, weight, and length, two measurements were made, and an average value was taken. The anthropometric measurements were converted to Weight for Height z-scores (WHZ) for determining the nutritional status of the 6 – 23-month-old children.

#### Data quality assurance

Data were collected by trained health extension workers. Before the commencement of the data collection, the research team provided detailed training to data collectors and supervisors on the objectives of the study, the data collection tools, and techniques. The training included both theoretical and practical sessions. After the completion of the training, trainees took a standardization test of anthropometric measurements. Those who scored passing marks were recruited as data collectors.

Besides, the data management team followed a real-time monitoring system to track data collection errors and provide timely corrective feedback to the data collectors. Range and consistency checks were built into the data entry system. Moreover, real-time GPS tracking of the point of data collection, online real-time checks, and feedback for missing values, inconsistencies, skip patterns, etc. were built into the software. Random spot checks were conducted by the research team members to validate the data collected by each data collector. Aggregate checks were conducted by comparing data collected against established existing population norms and gold standard data every month. Based on the above methods, a monthly appraisal of data collectors was conducted. Besides, quarterly quality audits were conducted by the research team.

### Data analysis strategies

The data collected were cleaned, coded, and analyzed using R version 4.4.1. Before being used in final analyses, raw anthropometric data were first entered into ENA software for calculation of the Z-scores.

Continuous variables were analyzed using means and standard deviations if normally distributed and median and interquartile range if not. For categorical variables, the chi-square was used to present frequencies and percentages. For the main analysis, first, differences in prevalence of acute malnutrition for each group were determined by subtracting the end line prevalence from baseline prevalence using the per protocol analysis. Difference -in-differences (DID) was used to compare the change in the prevalence of acute malnutrition for the treatment group to the change in the prevalence of acute malnutrition in the control group. The difference between these two changes was considered as the estimated effect of SQ – LNS on the burden of acute malnutrition.

### Ethical considerations

This interventional study was conducted in accordance with the Helsinki Declaration on Medical Research Involving Human Subjects. Ethical approval was obtained from the Institutional Review Board of the Tigray Health Research Institute (THRI/87/2023). Before the start of data collection, verbal informed consent were obtained from mothers or caregivers of children.

## Results

A total of 4,963 children in the intervention (SQ-LNS) group and 618 children in the control group were included in the analysis. The mean age of children was 13.95 (±5.27) months in the intervention group and 14.36 (±5.26) months in the control group. Similarly, the mean weight was 8.51 (±1.37) kg in the intervention group and 8.44 (±1.26) kg in the control group. The mean MUAC was also comparable between the two groups, measuring 13.17 (±0.96) cm in the intervention group and 13.19 (±0.83) cm in the control group. In terms of sex distribution, approximately half of the children were female in both groups (48.4% in the intervention group and 48.7% in the control group). The majority of children in both groups were born at term, although a slightly higher proportion of term births was observed in the intervention group. The prevalence of oedema was very low in both groups, with nearly all children having no oedema at 1st visit. However, a small difference was observed in mean height, with children in the control group having a slightly higher average height (73.51 ± 5.68 cm) compared to those in the intervention group (72.30 ± 5.84 cm) (Table 1).

**Table 1:** First visit characteristics of the 6 – 23-month-old children included in the SQ-LNS follow-up study in the post-conflict settings of Tigray, Northern Ethiopia.

| Variable | | Control<br>(Mean ± SD [95% CI]) | Intervention (SQ-LNS)<br>(Mean ± SD [95% CI]) |
| --- | --- | --- | --- |
| Age in months | | 14.36 ± 5.26 [13.95, 14.78] | 13.95 ± 5.27 [13.81, 14.10] |
| Weight in kg | | 8.44 ± 1.26 [8.34, 8.54] | 8.51 ± 1.37 [8.48, 8.55] |
| Height in cm | | 73.51 ± 5.68 [73.06, 73.96] | 72.30 ± 5.84 [72.14, 72.46] |
| MUAC in cm | | 13.19 ± 0.83 [13.12, 13.26] | 13.17 ± 0.96 [13.15, 13.20] |
| Sex | Female | 301 (48.7%) | 2401 (48.4%) |
|  | Male | 317 (51.3%) | 2562 (51.6%) |
| Birth status | Term | 575 (93.0%) | 4901 (98.8%) |
|  | Preterm | 34 (5.5%) | 61 (1.2%) |
|  | Do not know | 9 (1.5%) | 1 (0.0%) |
| Edema | Grade1 | 0 (0.0%) | 6 (0.1%) |
|  | No | 618 (100.0%) | 4957 (99.9%) |

### Frequency of measurements in the dynamic cohort

In this dynamic cohort, in which children entered at 6 months of age and exited at 24 months, the distribution of measurement frequency varied markedly between the control and intervention arms. A total of 1,393 and 6,752 children were enrolled into the control and intervention arms, respectively. Overall, 56.6% 33.5% of children in the control arm were measured four times or fewer and six or more times, respectively. In contrast, 56.6% of the intervention arm were measured six or more times, with a substantial 22.8% contributing ten, eleven, or twelve measurements (Table 2).

**Table 2:** Frequency of measurement among children in the dynamic cohort with age-based enrollment and exit in the SQ-LNS follow-up study in the post-conflict settings of Tigray, Northern Ethiopia.

| Measurement frequency<br>(Number of visits) | Number of children, n (%) |  |
| --- | --- | --- |
|  | Control, n (%) | Intervention, n (%) |
| 1 | 237 (17.0) | 573 (8.5) |
| 2 | 218 (15.6) | 586 (8.7) |
| 3 | 170 (12.2) | 584 (8.6) |
| 4 | 164 (11.8) | 583 (8.6) |
| 5 | 138 (9.9) | 606 (9.0) |
| 6 | 139 (10.0) | 520 (7.7) |
| 7 | 99 (7.1) | 553 (8.2) |
| 8 | 80 (5.7) | 598 (8.9) |
| 9 | 77 (5.5) | 607 (9.0) |
| 10 | 42 (3.0) | 685 (10.1) |
| 11 | 28 (2.0) | 601 (8.9) |
| 12 | 1 (0.1) | 256 (3.8) |
| Total | 1,393 (100) | 6,752 (100) |

### Impact of SQ-LNS on acute malnutrition Burden of acute malnutrition

Figure 2 shows the effect of the SQ-LNS intervention on the burden of global acute malnutrition (GAM) among the study children. The intervention group demonstrated a sustained reduction in GAM prevalence throughout the follow-up period, amounting to a total reduction of 17.6 percentage points (from 22.2% to 4.6%). The control group also showed improvement over time, but the decline was more gradual and less pronounced, representing a total reduction of 6.1 percentage points (from 20.11% to 14.0%) (Figure 2).

**Figure 2:**
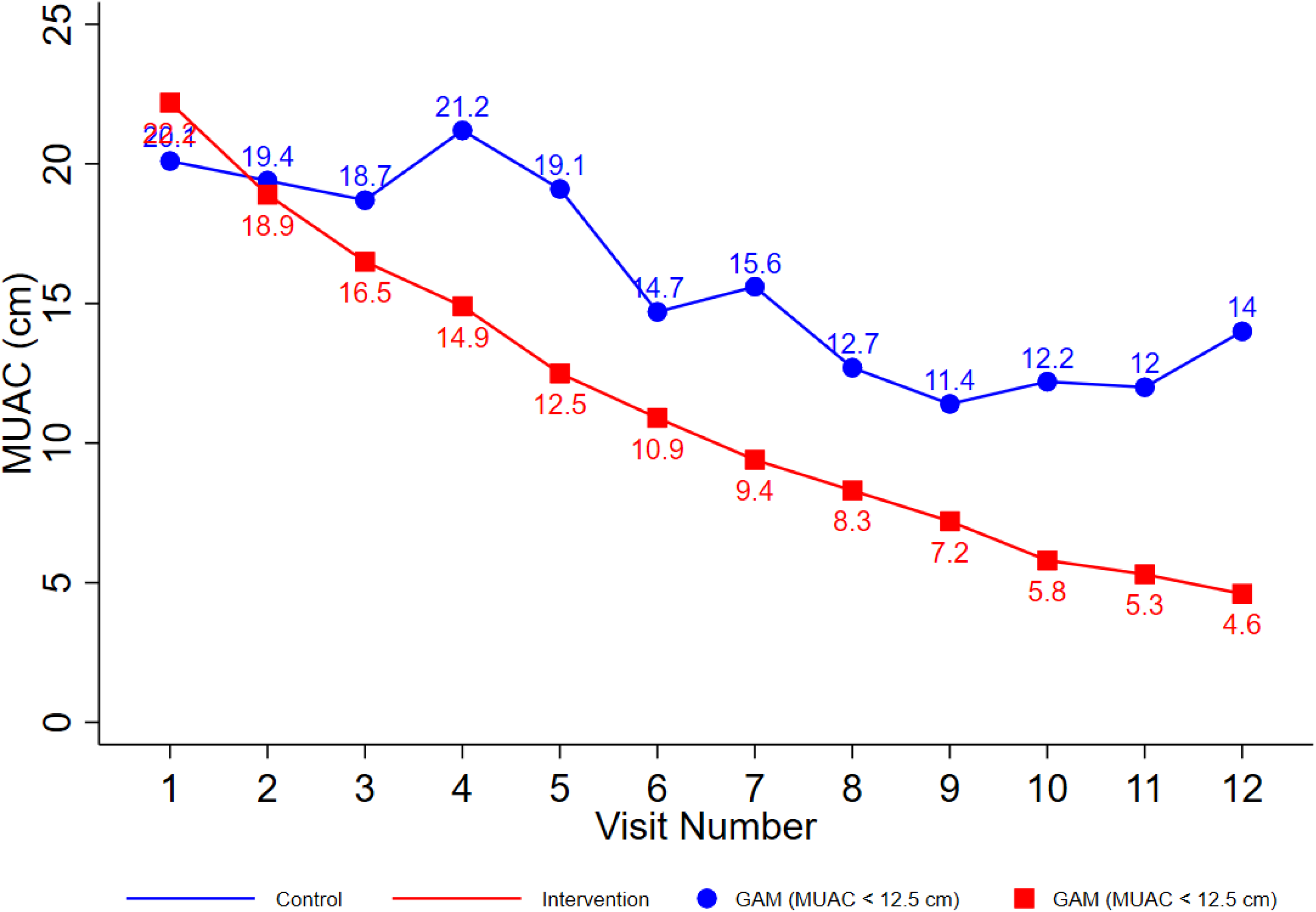
Impact of the daily provision of one sachet of SQ-LNS for six months on the burden of GAM (MUAC < 12.5 cm) among 6 - 23 months old children from the post conflict settings of Tigray, Ethiopia, 2024.

When the impact of the intervention is evaluated using WHZ, the values suggest a general decline in the intervention group over time, while control remains higher overall (Figure 3).

**Figure 3:**
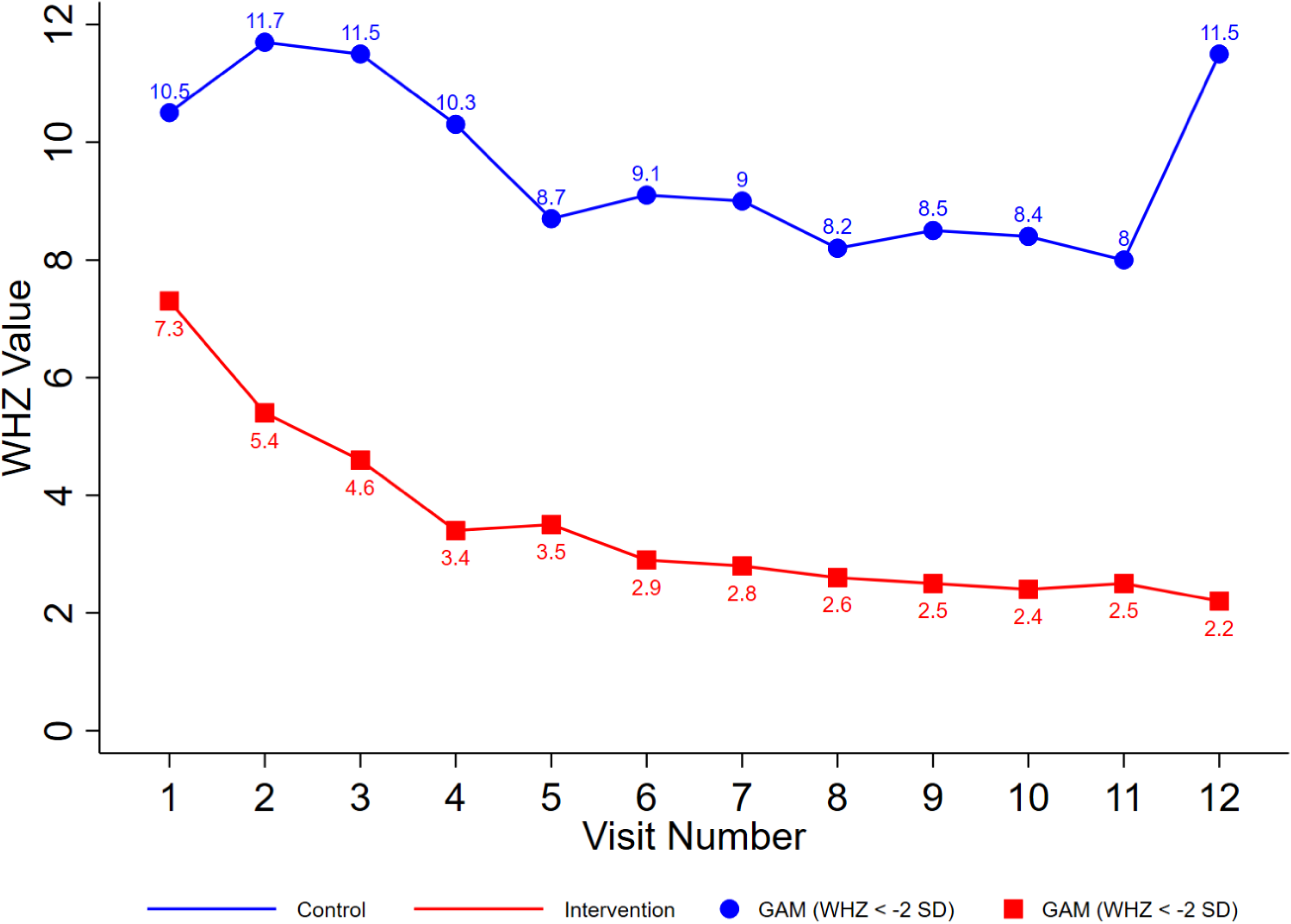
Impact of the daily provision of one sachet (20g) of SQ-LNS for six months on the burden of GAM (WHZ < -2 SD) among 6 - 23 months old children from the post conflict settings of Tigray, Ethiopia, 2024.

### Mean weight-for-height (WHZ) z scores

The provision of one SQ-LNS sachet daily to 6 – 23 months old children for six months had improved the mean weight-for-height z scores (WHZ) compared to the children from the control communities in Tigray, Ethiopia. The mean weight-for-height z-scores (WHZ) over the course of the study provide evidence of the rightward shift in nutritional status among children receiving SQ-LNS. The intervention group exhibited a consistent improvement in mean WHZ across the follow-up period, achieving a positive mean WHZ with a total gain of +0.72 z-score units (from - 0.35 to +0.37) compared to the control group with a total gain of +0.16 (from -0.79 to -0.63) (Figure 4).

**Figure 4:**
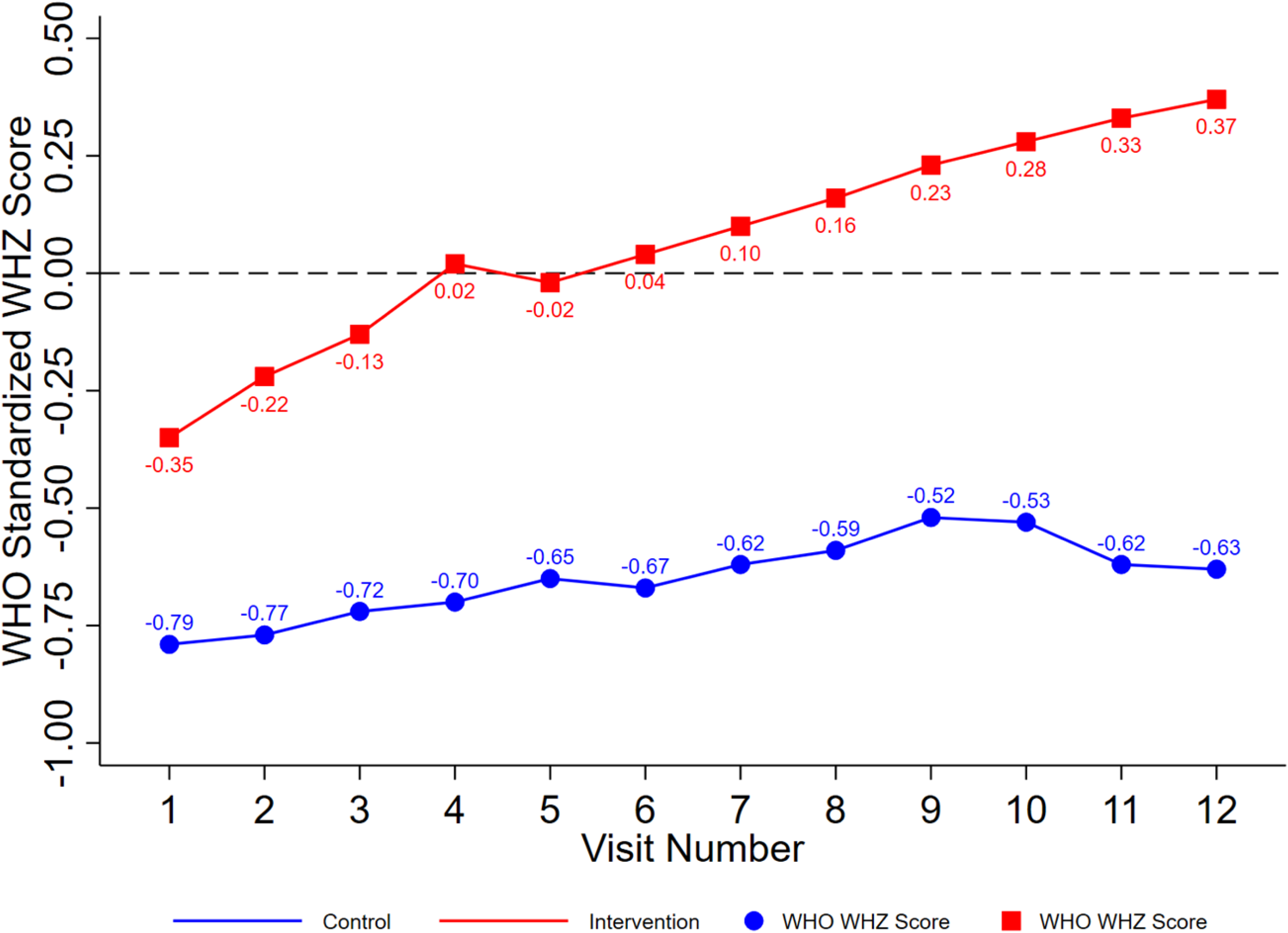
Illustration of the impact of one sachet of SQ-LNS supplementation for six months on the WHZ scores of 6 – 23 months old children from the post-conflict communities in Tigray, Ethiopia, 2024.

The SQLNS intervention has pushed the ’floor’ of the distribution to the right and caused a clear rightward shift in the distribution of WHZ scores. The entire bell curve of nutritional status has moved away from the risk of wasting and toward the healthy standard (Figure 5).

**Figure 5:**
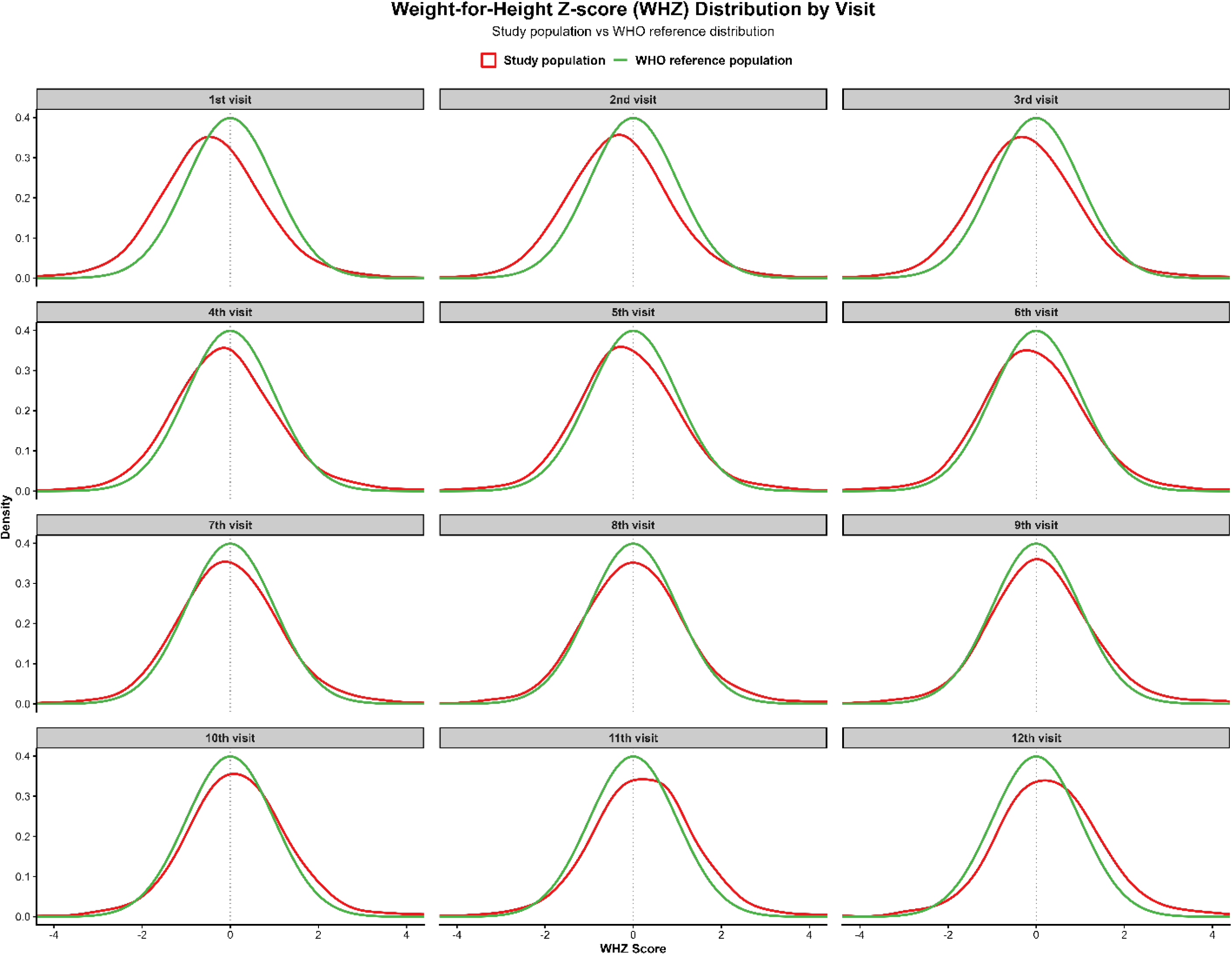
Figure 4: Rightward shift in WHZ scores following SQLNS intervention among 6 – 23 months old children from post-conflict settings of Tigray, Ethiopia.

### Difference in difference (DiD) Burden of malnutrition

The difference-in-differences (DiD) estimation showed that the provision of SQ-LNS reduced the prevalence of acute malnutrition (low weight-for-height) among children compared with those in the control communities (Table 3). At the beginning, the intervention group had a prevalence of GAM that was 3.2 percentage points (ppt) lower than the control group. Over the course of the study, the control group saw a natural reduction of 1 ppt, and the intervention group experienced a much sharper decline of 5.1 ppt. After accounting for the reduction seen in the control group, the net effect of the intervention (the DiD estimate) was a net reduction of 4.1 percentage points in GAM prevalence as defined by WHZ. When the GAM prevalence was defined by MUAC, it dropped by 17.6 ppt in the intervention group and by 6.1ppt in the control group, with a net reduction of 11.5 ppt in GAM prevalence based on MUAC (Table 3).

**Table 3:** Difference in difference estimation of the impact of SQ-LNS on the prevalence of acute malnutrition among 6 – 23 months old children from the post conflict communities in Tigray, Ethiopia, 2024.

| Changes |  | Indicators |  |
| --- | --- | --- | --- |
|  |  | GAM (WHZ < -2 z scores) | GAM (MUAC < 12.5 cm) |
| Baseline (first visit) difference | Intervention – Control | 7.3 – 10.5 = - 3.2 ppt | 22.2 – 20.1 = +2.1 ppt |
| Control group change | 12<sup>th</sup> visit – first visit | 11.5 – 10.5 = 1 ppt | 14.0 – 20.1 = -6.1 ppt |
| Intervention group change | 12<sup>th</sup> visit – first visit | 2.2 - 7.3 = - 5.1 ppt | 4.6 - 22.2 = -17.6 ppt |
| DiD Estimate<br>(Net effect) | 12<sup>th</sup> visit – first visit | -5.1 - (1) = -4.1 ppt | -17.6 – (-6.1) = -11.5 ppt |

### Mean WHZ z scores and MUAC measurements

The difference in difference (or “double difference”) estimation, or the difference in average outcome in the SQ-LNS treatment group before and after treatment, minus the difference in average outcome in the control group before and after treatment, revealed an increase in weight- for-height z scores and MUAC in the communities where the children were provided with SQ-LNS (Table 4). The Mean WHZ scores of the intervention group increased from a baseline of negative 0.35 at the first visit to an average of positive 0.37 by the 12th visit, representing a substantial overall gain of 0.72 points. In contrast, the control group showed only modest improvement, moving from negative 0.79 to an average of negative 0.63 by the 12th visit, for a net gain of just 0.16 points. Similarly, the intervention group’s mean MUAC increased from 131.00 mm at the first visit to an average of 138.15 mm at the 12th visit with a total gain of 7.15 mm. The control group’s mean MUAC increased marginally from 132.10 mm to 134.22 mm at the 12th visit, resulting in a total gain of only 2.12 mm. The net gain from the DiD estimate for mean WHZ scores and MUAC measurements was 0.56 z scores and 5.03mm, respectively (Table 4).

**Table 4:**
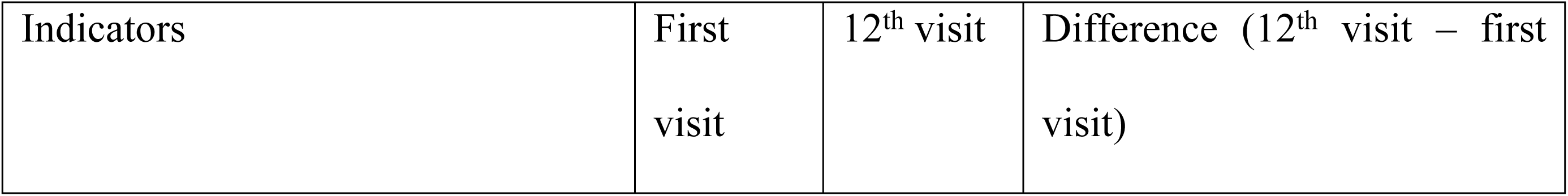
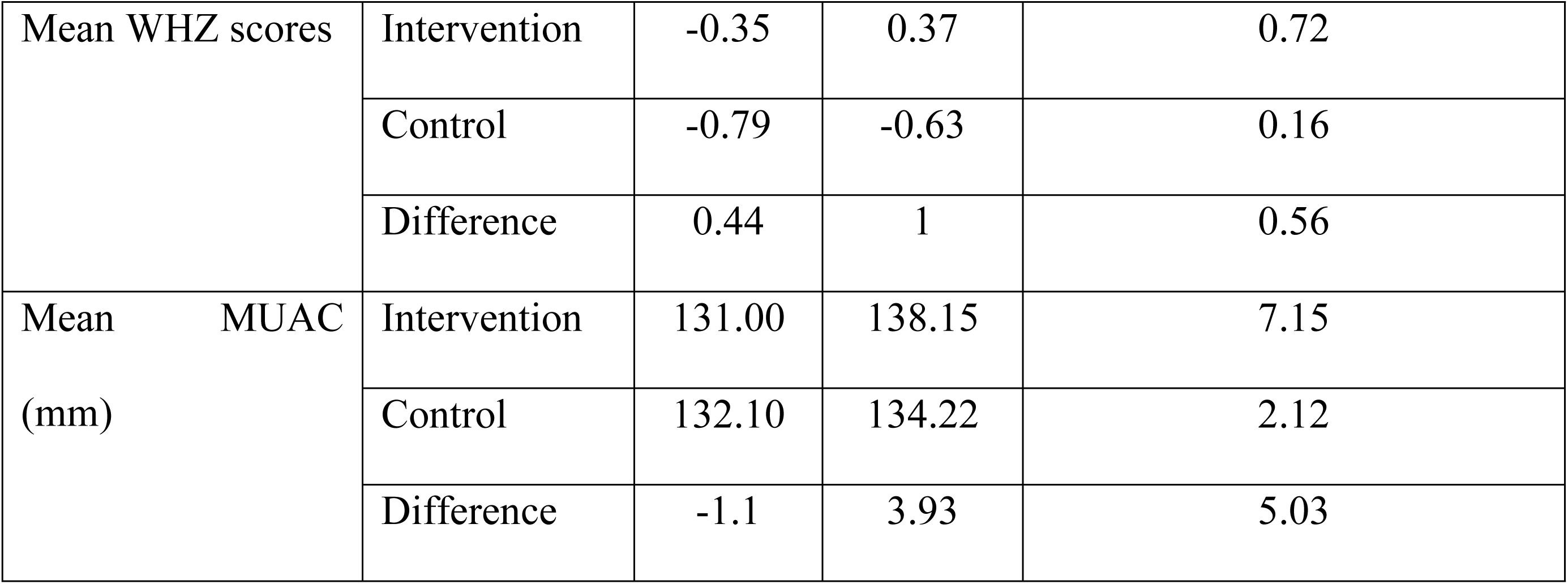
Difference in difference estimation of the impact of SQ-LNS on the mean z scores and MUAC measurements among 6 – 23 months old children from the post conflict communities in Tigray, Ethiopia, 2024.

| Indicators | First<br>visit | 12<sup>th</sup> visit | Difference (12<sup>th</sup> visit – first<br>visit) |
| --- | --- | --- | --- |

|  |  |  |  |  |
| --- | --- | --- | --- | --- |
| Mean WHZ scores | Intervention | -0.35 | 0.37 | 0.72 |
|  | Control | -0.79 | -0.63 | 0.16 |
|  | Difference | 0.44 | 1 | 0.56 |
| Mean MUAC<br>(mm) | Intervention | 131.00 | 138.15 | 7.15 |
|  | Control | 132.10 | 134.22 | 2.12 |
|  | Difference | -1.1 | 3.93 | 5.03 |

## Discussion

In the present study, we sought to assess the effect of the provision of SQ-LNS on the protective action of acute malnutrition among 6 – 23 months old children compared to the existing nutrition interventions in the control communities. Findings of this study suggest that SQ-LNS is an effective intervention for reducing the occurrence of acute malnutrition and bringing a rightward shift of the entire population distribution in 6 – 23 months old children from post-conflict settings. Daily supplementation of SQ-LNS for six months resulted in a decrease in the burden of GAM (based on MUAC) from 22.2% to 4.6% compared to the GAM reduction from 20.1% to 14.0% in the control woredas. Similarly, the burden of GAM (based on WHZ scores) decreased from 7.3 to 2.2% in the SQ-LNS and from 10.5 to 11.5% in the control communities. The increase in the mean WHZ score from negative to positive (-0.35 to +0.37), and a net mean WHZ score gain of +0.56 z scores (DiD estimate) shows that the nutritional status of the average child in the intervention group shifted into a healthier range. This “rightward shift” of the entire population distribution is a key piece of evidence supporting the argument that the benefits of SQ-LNS go “beyond prevention.”

The 11.5 percentage point reduction (-17.6% vs -6.1%) based on MUAC and 4.1 percentage point reduction (-5.1% vs 1%) based on WHZ observed in the intervention group provides compelling evidence of the protective effect of SQ-LNS against acute malnutrition among 6 – 23 months old children from post conflict settings. Furthermore, the uninterrupted decline in the intervention group as compared with the fluctuating pattern in the control group suggests that the SQ-LNS intervention provided sustained protection against acute malnutrition throughout the study period. The follow-up data further demonstrated that while secular trends or seasonal factors may have contributed to some improvement in both groups (as evidenced by the control group’s decline from 20.1% to 14.0%), the SQ-LNS intervention accelerated the reduction of the burden of acute malnutrition in the intervention group. The findings are in line with the meta-analysis of individual participant data from 14 randomized controlled trials of SQ-LNSs provided to children 6–24 months of age, which reported a decrease of wasting by 14 - 18% [5]. Similarly, the meta-analysis of individual participant data from 14 randomized controlled trials of SQ-LNSs provided to children 6–24 months of age led to a relative reduction of 31% in severe wasting at end line [16]. Lipid-based nutrient supplements given with complementary foods to infants and young children 6 to 23 months of age reduced moderate wasting by 18% [7]. Another systematic review of 17 studies on the provision of SQ-LNS to children aged 6–23 months reduced moderate wasting by 17% [17], suggesting that SQ-LNS is an effective intervention to address acute malnutrition among 6 – 23 months old children. Acute malnutrition (wasting) is caused by a combination of inadequate dietary intake (not enough calories and nutrients) and frequent infections (which decrease appetite and increase nutrient loss). The justification lies in its direct action on the two primary causes of acute malnutrition. The SQ-LNS directly addresses the causes of acute malnutrition by filling the nutrient gaps through the provision of high-quality protein, essential fats, and a full spectrum of vitamins and minerals (iron, zinc, vitamin A, etc.) that are often missing from the complementary foods given to young children in resource-poor settings. The fat-rich paste of SQ-LNS provides a concentrated source of calories, helping children meet their high energy needs for growth and activity. The fortified and comprehensive package of vitamins and minerals prevents micronutrient deficiencies like zinc, iron, and vitamin A and maintains a robust immune system. A stronger immune system means fewer and less severe infections (like diarrhea and pneumonia), which are major precipitants of acute malnutrition. Moreover, the SQ-LNS ensures the body has the necessary building blocks, and helps maintain muscle and fat mass and eventually prevents acute malnutrition or wasting among the 6 – 23 months old children from the post conflict settings of Tigray, Ethiopia. The country carries a severe burden of acute malnutrition, with a prevalence of 11% among children under five, a rate that has shown no sign of reduction for two decades [4, 12–14, 18]. To meet the 2030 SDG target of reducing wasting to below 5%, the current Annual Average Reduction Rate (AARR) in Ethiopia must increase dramatically from 0.1% to approximately 5.4% [19], underscoring a pressing need for additional, evidence-based preventive strategies. Scaling up the distribution and access to SQLNS for young children presents a critical opportunity to bridge this gap. As a proven intervention for preventing acute malnutrition, a large-scale SQLNS program could directly contribute to increasing the AARR, thereby accelerating progress toward the SDG target and protecting a generation of Ethiopian children from the lifelong consequences of wasting.

SQ-LNS has improved the mean WHZ of 6 – 23-month-old children with a total gain of +0.72 z-score units, from -0.35 to +0.37. The improvement in the mean WHZ in the intervention group in the present study reveals that the entire distribution of nutritional status has shifted rightwards (crossing from a negative z score to a positive z score), indicating that even children who were not classified as malnourished experienced measurable gains in weight-for-height. The intervention did not merely rescue the children with acute malnutrition, but it shifted the curve for the entire population. This explains why the intervention group achieved a GAM (WHZ) prevalence of 1.3% within the WHO global target to achieve the 2025 target to reduce and maintain child wasting to less than 5% [20] while simultaneously achieving a positive mean WHZ, a sign of population-level nutritional resilience [21]. The control group, by comparison, shows no decline in prevalence (11.5 to 10.5%) and remained with a negative mean WHZ (-0.63), indicating that the average child was still lighter than the WHO standard. Thus, both the reduction in the prevalence of acute malnutrition and the rightward shift in mean WHZ revealed compelling evidence of both curative impact (reducing severe cases) and population-level nutritional enhancement (lifting the average child) through the provision of SQLNS for a period of six months to 6 – 23 months old children. The present findings demonstrated that SQ-LNS does more than just treat malnutrition; rather, it builds broader nutritional resilience in communities with the highest burden of acute malnutrition among 6 – 23 months old children. This indicates that SQ-LNS conferred benefits that extended across the full spectrum of children, not just those at the highest risk of acute malnutrition, for children in post-conflict settings.

The DiD analysis reveals that SQ-LNS had a positive and meaningful impact on both indicators (WHZ and MUAC), but the nature of the effect differed. For WHZ, the intervention not only improved the mean but also closed the gap with the WHO standard population, achieving a positive score. For MUAC, the intervention accelerated growth in arm circumference, with a net gain of 5.03 mm attributable to SQ-LNS. The consistency of the positive DiD estimates across both indicators provides evidence that SQ-LNS supplementation improved nutritional status, regardless of which anthropometric indicator is used. The net effect of +0.56 z-score units for WHZ and +5.03 mm for MUAC represents the true impact of the intervention after accounting for baseline differences and secular trends. The DiD estimate of +0.56 z-score units quantifies the extent to which SQ-LNS contributed to building the nutritional resilience. The MUAC DiD estimate of +5.03 mm reflects muscle mass and subcutaneous fat stores that can be mobilized during periods of physiological stress. A net gain of 5.03 mm in arm circumference attributable to SQ-LNS represents a meaningful increase in these reserve tissues. Children with greater arm circumference have more metabolic capital to draw upon during illness or food scarcity, making them less likely to experience rapid deterioration into severe acute malnutrition. The fact that the intervention group started with slightly lower MUAC at baseline (131.00 mm vs. 132.10 mm) but ended with higher MUAC by the final visit (138.15 mm vs. 134.22 mm) illustrates the power of the SQ-LNS intervention to not only close but reverse an initial disadvantage. The dual effect of gaining external growth (weight-for-height) and internal reserves (arm circumference) via SQLNS has important implications for post-conflict settings, where populations often face recurrent stressors [22]. The DiD analysis provides rigorous quantitative evidence that the SQ-LNS intervention produced a rightward shift in the distribution of nutritional status and hence builds a foundation of nutrition resilience that can protect children in post conflict settings against future episodes of malnutrition. The present findings support the scale-up of SQ-LNS interventions not only as a treatment for acute malnutrition but as a strategy for protecting vulnerable children from post conflict settings against nutritional deprivation.

The present study has limitations that should be considered when interpreting the findings. The non-randomized design is one of the limitations of the study. Without random assignment, the potential for selection bias cannot be eliminated. Although we employed DiD analysis to account for time-invariant differences between groups, unmeasured time-varying confounders may have influenced the results. While DiD methodology addresses fixed differences, it cannot fully adjust for differential trajectories that might have occurred even in the absence of the intervention. Secondly, the difference in sample size between the intervention at first visit (n=4993) and control (n=618) groups may have potentially masked heterogeneity in intervention effects. However, the consistency of findings across multiple indicators and time points strengthens confidence in the results. Third, the generalizability of the findings may be limited. The study was conducted in a specific post-conflict context with its unique characteristics. Extrapolation to other settings should be done cautiously. Despite these limitations, the study has important strengths, including the large sample size, the extended follow-up period with multiple visits, the use of rigorous DiD methodology, and the examination of both prevalence and distributional outcomes. The consistency of findings across different analytical approaches and the magnitude of the observed effects lend credibility to the conclusions of the present findings.

## Conclusions and recommendations

In conclusion, our study findings demonstrated that provision of the 20 g SQ-LNS to 6 – 23 months old children daily for six months reduced the burden of acute malnutrition and improved the WHZ scores among 6 – 23 months old children from Tigray, Ethiopia. The SQ-LNS intervention not only reduced the prevalence of acute malnutrition but also shifted the entire distribution of weight-for-height rightward, building population-level nutritional resilience. We recommend that policymakers and program designers consider SQ-LNS as a hybrid intervention capable of simultaneously preventing acute malnutrition and building nutritional capital that protects against future deterioration among 6 – 23 months old children from post-conflict settings.

## Data Availability

Authors recognize that we are required to make fully available and without restriction all data underlying their findings.

## Acknowledgments

We would like to acknowledge UNICEF for funding and close follow-up of the trial implementation and the Regional Health Bureau (RHB), Tigray Development Association (TDA) and Mums for Mums (MfMs) for the implementation of the SQ-LNS trial in Tigray, Ethiopia. We also thank the families and communities who participated in the SQ-LNS trial, and all of the members of the SQ-LNS team from the implementing partners, namely RHB, TDA and MfMs. Special thanks go to the field supervisors of TDA and MfMs for their close follow-up with the trial implementation and project management. The support from the Health Extension Workers and Health Care Professionals during the collection of the anthropometric data and the home visits is duly acknowledged.

